# Fracture risk among stroke survivors according to post-stroke disability status and stroke type

**DOI:** 10.1101/2023.12.19.23300259

**Authors:** Dagyeong Lee, In Young Cho, Won Hyuk Chang, Jung Eun Yoo, Hea Lim Choi, Jun Hee Park, Dong Wook Shin, Kyungdo Han

**Affiliations:** Department of Family Medicine/Supportive Care Center, Samsung Medical Center, Sungkyunkwan University School of Medicine, Seoul, Republic of Korea; Department of Clinical Research Design & Evaluation, Samsung Advanced Institute for Health Science & Technology, Sungkyunkwan University, Seoul, Republic of Korea; Department of Physical & Rehabilitation Medicine, Samsung Medical Center, Sungkyunkwan University School of Medicine, Seoul, Republic of Korea; Department of Family Medicine, Healthcare System Gangnam Center, Seoul National University Hospital, Seoul, Republic of Korea; Department of Family Medicine/Executive Healthcare Clinic, Severance Hospital, Yonsei University College of Medicine, Seoul, Republic of Korea; Department of Medicine, Sungkyunkwan University School of Medicine, Seoul, Republic of Korea; Department of Statistics and Actuarial Science, Soongsil University, Seoul, Republic of Korea

**Keywords:** stroke, fracture, disability, cohort

## Abstract

**Background:** Stroke survivors face physical and cognitive challenges, including impaired coordination and balance, which can lead to an increased dependency and a higher risk of falls. We aimed to investigate the impact of post-stroke disability status and stroke type on the risk of fracture at various sites compared to a matched comparison group.

**Method:** This retrospective cohort study used data from the Korean National Health Insurance System database (2010-2018) and included a total of 223,358 stroke patients and a 1:1 matched comparison group. Stroke survivors were grouped based on the presence and severity of their post-stroke disability and stroke type. The primary outcome was the incidence of newly diagnosed fracture. Cox proportional hazard regression analyses were used to calculate the hazard ratios of fractures after adjusting for potential confounders.

**Results:** Stroke survivors had an increased risk of overall fractures compared to the matched comparison group (adjusted hazard ratio [aHR] 1.40, 95% confidence interval [CI] 1.37-1.43). Specifically, the risk of hip fractures was even greater for stroke survivors: aHR 2.42, 95% CI 2.30-2.55. The risk of vertebral fractures (aHR 1.29, 95% CI 1.25-1.34) and other fractures (aHR 1.19, 95% CI 1.15-1.23) also was higher than that of the control group. The risk of hip fractures was highest among stroke survivors with severe post-stroke disability (aHR 4.82, 95% CI 4.28-5.42), while the risk of vertebral or other fractures was highest among those with mild post-stroke disability. There was no significant difference in fracture risk between hemorrhagic and ischemic stroke survivors when stratified by disability status.

**Conclusion:** Our findings showed an increased risk of subsequent fractures among stroke survivors, particularly those with post-stroke disability and for hip fracture. Bone health assessment and treatment should be emphasized as an essential part of stroke management.

## Introduction

According to the Global Burden of Diseases 2019, over 12.2 million new stroke events occur annually^1^. Stroke is the second most prevalent cause of death and the third most significant contributor to disability-adjusted life years worldwide^2^. Stroke can have a significant impact on both physical and cognitive abilities^3^, resulting in coordination and balance impairments^4^. Muscle weakness, immobility, and dependency following stroke can also lead to physical impairment associated with decreased quality of life and higher mortality^5^. Moreover, stroke survivors face an elevated risk of falls^6^.

Although these factors suggest that stroke survivors are at increased risk of fractures, previous studies reported heterogeneous results (**Table S1**)^7–17^. For example, hip fracture risk in stroke survivors compared to a non-stroke group varied across studies, with reported relative risk (RR) of 1.4 to 2.4 in a Taiwanese study^12^, RR 11.75 in a Swedish study^13^, and no evidence of increased risk in a US study (adjusted hazard ratio [aHR] 1.1, 95% confidence interval [CI] 0.6-2.1)^17^. In addition, previous studies primarily focused on hip^9,12,13,15,16^ or overall fractures^7,9,13^, and direct comparisons among different fracture sites were rarely reported^7,11^. The influence of severity of disability on fracture risk remains uncertain. A US study^9^ found that post-stroke women with severe disability were at increased risk of hip fractures (aHR 2.1, 95% CI 1.4-3.2), but a Taiwanese study^7^ showed an inverse relationship between stroke severity and overall fracture (aHR for moderate vs mild stroke, 0.88; 95% CI 0.81-0.96, aHR for severe vs mild stroke, 0.39; 95% CI 0.34-0.44). The effect of stroke type on fracture risk has also been inconsistent; some studies indicated a higher fracture risk in hemorrhagic stroke compared to ischemic stroke^18,19^, but others reported no significant difference^7^. Moreover, most studies on fracture among stroke survivors failed to differentiate by stroke type^8,12,14–17^.

In this retrospective cohort study, we aimed to investigate the risk of fractures among stroke survivors compared to a non-stroke population. We also investigated the impacts of post-stroke disability and stroke type on fracture at various sites. We hypothesized that stroke survivors would have a higher risk of fractures compared to the control group, with the risk varying by fracture site and type of stroke. Additionally, we expected that the severity of post-stroke disability would correlate with an increased fracture risk.

## Methods

### 1. Data source

This study used the Korean National Health Insurance Service (KNHIS) database, which includes diagnoses and prescriptions for the entire Korean population^20^ (further details in **Supplementary Methods**). Under the KNHIS, employees and all people aged 40 or above are eligible for general health screening programs^21^. The KNHIS hence collects and manages information on demographic factors, anthropometric measurements, health behaviors, and laboratory results^20^. This database is a valuable resource for clinical and public health research^21^.

### 2. Study population

We identified a total of 800,646 individuals who experienced a stroke between January 1, 2010, and December 31, 2018. The definition of stroke was determined by ICD-10 codes (**Table S2**) from hospitalization records, along with the presence of claims for brain resonance imaging or brain CT scan^22^. We matched stroke survivors with a comparison group who had no previous stroke event in a 1:1 ratio based on age and sex. The control subjects were assigned an index date, which aligned with the date of stroke diagnosis for their respective matched patients.

Among the selected stroke (n=800,646) and comparison (n=800,646) groups, we included those who underwent a national health check-up within two years of the index year (n=465,257 and n=391,878) to retrieve baseline information on health behaviors and comorbidities. We excluded individuals who were younger than 40 years (n=6,719 and n=8,300) to analyze a population at risk for fractures, our outcome of interest. We also excluded participants with a previous diagnosis of fracture (n=62,858 and 59,785) or missing data on covariates (n=13,036 and 13,713) and applied a 1-year lag (n=29,418 and 4,809). The 1-year lag was introduced to examine the risk of fracture after the stroke has stabilized and to exclude fractures that may occur during the acute stroke phase. In total, 223,358 were included in the stroke group and 322,161 in the comparison group (**Figure 1**). This study was approved by the Institutional Review Board of Samsung Medical Center (IRB No. 2020-12-068).

**Figure 1.**
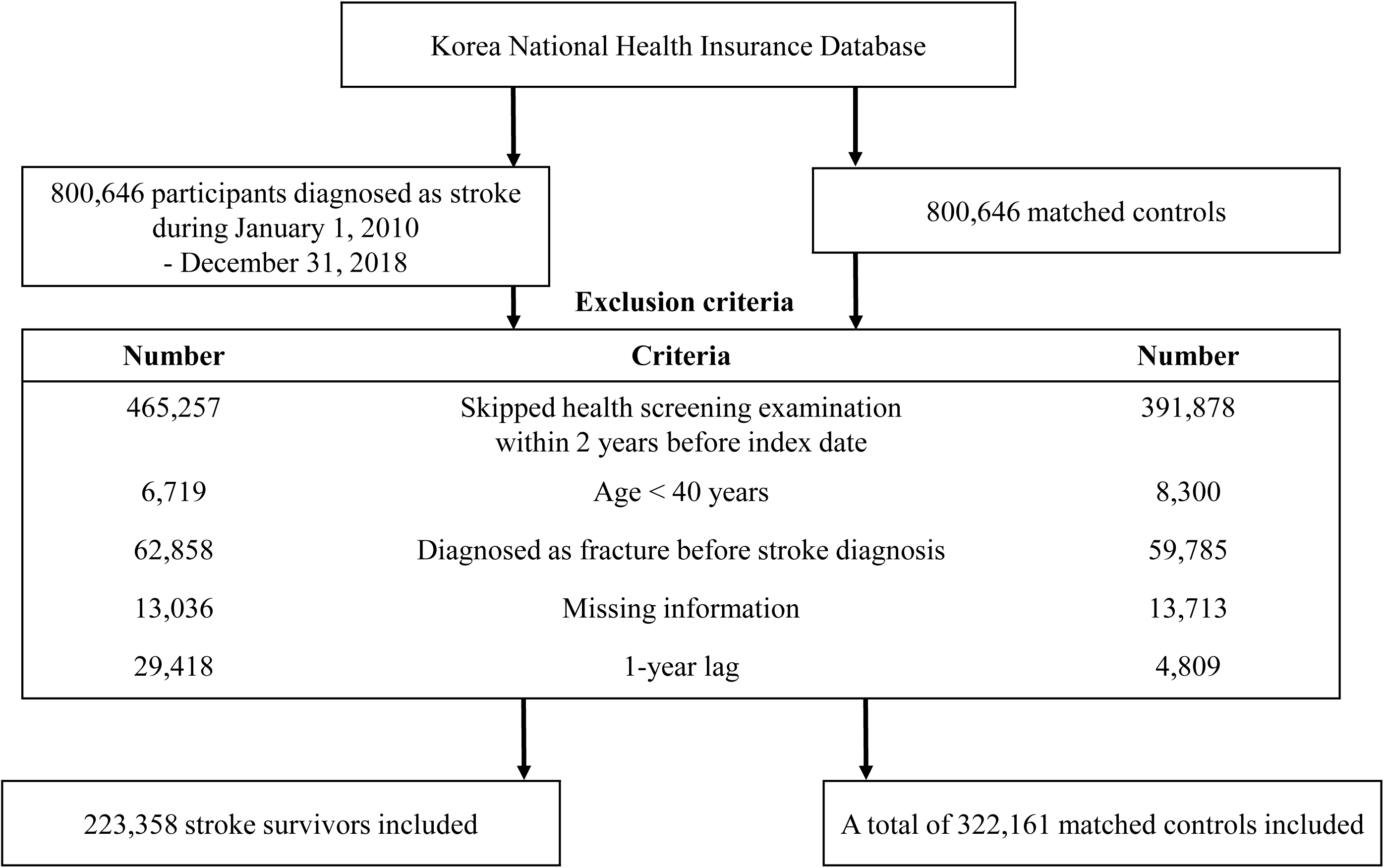
Flowchart of the study population

### 3. Definition of disability and its severity caused by stroke

We used data from Korea’s National Disability Registration System (KNDRS) to determine post-stroke disability status and severity (further details in **Supplementary Methods**). Disability status and severity information from the KNDRS can be considered reliable and accurate. Grade 1 (most severe) disability from brain injury was defined by the requirement of total assistance from others, whereas grade 6 (least severe) disability was defined by the performance of ordinary activities independently according to predefined eligibility criteria (**Table S3**)^23^. For the analyses, we classified grades 1 to 3 as severe disability and grades 4 to 6 as mild disability.

### 4. Study outcomes and follow-up

The primary outcome of this study was a newly diagnosed fracture, determined by ICD-10 codes^24^ (**Table S4**). Hip fracture was defined with the relevant diagnosis codes; vertebral and other fractures were defined by two or more outpatient visits with the relevant diagnosis codes within a 12-month period. “Overall fracture” refers to the occurrence of at least one of the above-mentioned fractures. The study population was followed until the occurrence of a new fracture, censor date, or the end of the study period (December 31, 2018), whichever came first.

### 5. Covariates

Sociodemographic information such as age, sex, residential area, and income level was obtained from the KNHIS database. Data on lifestyle behaviors (smoking, alcohol consumption, and physical activity), anthropometric measurements, and laboratory test results were extracted from the general health screening examinations.

Comorbidities of the participants were identified using claims and prescription information prior to the index date (further details in **Supplementary Methods**).

### 6. Statistical analysis

Descriptive analysis was conducted using mean ± standard deviation (SD) for continuous variables and number (percentage) for categorical variables. Cox proportional hazards regression analysis was performed to calculate the hazard ratio (HR) and 95% CI for fracture risk among stroke survivors compared to the matched comparison group, with adjustment for potential confounders during years of follow-up. Model 1 was unadjusted, and model 2 was adjusted for age (continuous), sex, area of residence (urban, rural), insulin use, diabetes mellitus (DM), hypertension, dyslipidemia, smoking (none, ex, current), alcohol consumption (non, mild, heavy), and body mass index (BMI) (continuous). In addition, individuals with stroke were categorized into three groups: (1) stroke without disability, (2) stroke with mild disability (grades 4–6), and (3) stroke with severe disability (grades 1–3). We conducted Kaplan-Meier analyses to demonstrate the cumulative incidence probabilities of fracture.

All statistical analyses were conducted using SAS version 9.4 (SAS Institute Inc., Cary, NC, USA). Two-sided P-values <0.05 were considered statistically significant.

## Results

### 1. Baseline characteristics

Among 223,358 stroke survivors, the mean age was 64.8 ± 10.9 years, and 61.2% were male (**Table 1**). Stroke survivors were more likely to reside in rural areas and had lower incomes than the control group. Compared to the control group, a higher percentage of stroke survivors were current smokers (26.1% versus 17.6%), heavy drinkers (8.3% versus 6.7%), and had a higher BMI. Stroke survivors were more likely to have higher Charlson Comorbidity Index scores, and a higher prevalence of DM, hypertension, and dyslipidemia.

**Table 1.** Baseline characteristics of the study population.

|  | Study population |  | P-value | Stroke survivors |  | P-value |
| --- | --- | --- | --- | --- | --- | --- |
|  | Matched controls<br>(N= 322,161) | Stroke survivors<br>(N= 223,358) |  | Without post-stroke<br>disability<br>(N= 196,426) | With post-stroke<br>disability<br>(N= 26,932) |  |
| Mean age, years | 65.2 ± 11.2 | 64.8 ± 10.9 | <0.001 | 64.6 ± 11.0 | 66.5 ± 10.0 | <0.001 |
| Sex, male | 197,357 (61.3) | 136,623 (61.2) | 0.5 | 19,400 (60.8) | 17,223 (64.0) | <0.001 |
| Smoking |  |  | <0.001 |  |  | <0.001 |
| Non-smoker | 192,161 (59.7) | 124,483 (55.7) |  | 109,232 (55.6) | 15,251 (56.6) |  |
| Ex-smoker | 73,321 (22.8) | 40,633 (18.2) |  | 35,687 (18.2) | 4,946 (18.4) |  |
| Current smoker | 56,679 (17.6) | 58,242 (26.1) |  | 51,507 (26.2) | 6,735 (25.0) |  |
| Alcohol consumption |  |  | <0.001 |  |  | <0.001 |
| Non | 198,336 (61.6) | 136,167 (61.0) |  | 118,718 (60.4) | 17,449 (64.8) |  |
| Mild | 102,170 (31.7) | 68,701 (30.8) |  | 61,347 (31.2) | 7,354 (27.3) |  |
| Heavy | 21,655 (6.7) | 18,490 (8.3) |  | 51,507 (26.2) | 6,735 (25.0) |  |
| Regular physical activity | 72,905 (22.6) | 42,941 (19.2) | <0.001 | 38,147 (19.42) | 4,794 (17.8) | <0.001 |
| Anthropometrics |  |  |  |  |  |  |
| Body mass index, kg/m<sup>2</sup> | 24.1 ± 3.1 | 24.3 ± 3.2 | <0.001 | 24.3 ± 3.1 | 24.6 ± 3.3 | <0.001 |
| Waist circumference, cm | 83.1 ± 8.5 | 84.0 ± 8.6 | <0.001 | 83.8 ± 8.6 | 85.1 ± 8.7 | <0.001 |
| Systolic BP, mmHg | 127.0 ± 15.3 | 131.8 ± 17.4 | <0.001 | 131.7 ± 17.34 | 132.5 ± 17.4 | <0.001 |
| Diastolic BP, mmHg | 77.3 ± 9.8 | 80.2 ± 11.3 | <0.001 | 80.2 ± 11.3 | 80.3 ± 11.2 | <0.001 |
| Comorbidity |  |  |  |  |  |  |
| Hypertension | 164,380 (51.0) | 171,966 (77.0) | <0.001 | 149,710 (76.2) | 22,256 (82.6) | <0.001 |
| Diabetes Mellitus | 60,105 (18.7) | 67,565 (30.3) | <0.001 | 58,293 (29.7) | 9,272 (34.4) | <0.001 |
| Dyslipidemia | 114,553 (35.6) | 158,225 (70.8) | <0.001 | 139,603 (71.1) | 18,622 (69.1) | <0.001 |
| Laboratory findings |  |  |  |  |  |  |
| Glucose, fasting, mg/dL | 103.8 ± 26.2 | 110.6 ± 37.8 | <0.001 | 110.4 ± 37.7 | 132.5 ± 17.4 | <0.001 |
| eGFR, mL/min/1.73 m<sup>2</sup> | 85.0 ± 42.4 | 83.7 ± 40.4 | <0.001 | 83.8 ± 40.4 | 83.3 ± 40.4 | <0.001 |
| Total cholesterol, mg/dL | 194.5 ± 38.3 | 198.2 ± 40.8 | <0.001 | 198.5 ± 40.8 | 195.8 ± 41.3 | <0.001 |
| Urban residency | 141,212 (43.8) | 89,610 (40.1) | <0.001 | 79,442 (40.4) | 101,68 (37.7) | <0.001 |
| Income of lowest 25% | 61,013 (19.0) | 48,564 (21.7) | <0.001 | 41,902 (21.3) | 6,662 (24.7) | <0.001 |
| Charlson comorbidity index | 1.8 ± 1.9 | 4.3 ± 2.4 | <0.001 | 4.3 ± 2.3 | 4.8 ± 2.5 | <0.001 |
| Follow-up duration, years | 4.2 ± 2.5 | 3.7 ± 2.5 | <0.001 | 3.7 ± 2.5 | 3.7 ± 2.5 | <0.001 |

Data are expressed as mean ± standard deviation or number (%); BP, blood pressure; eGFR, estimated glomerular filtration rate.

Among 26,932 stroke survivors with post-stroke disability, the mean age was 66.5 ± 10.0 years, and 64.0% were male. Stroke survivors with post-stroke disability had lower incomes and a higher BMI than those without post-stroke disability. Stroke survivors with disability had higher Charlson Comorbidity Index scores than those without post-stroke disability, and a higher prevalence of DM and hypertension.

### 2. Risk of subsequent fracture among people with stroke according to post-stroke disability

The mean ± SD follow-up period for stroke survivors was 3.7 ± 2.5 years, with an average 2.7 ± 2.0 years between stroke and new-onset fracture. The comparison group had a mean ± SD follow-up period of 4.2 ± 2.5 years. The Kaplan-Meier curves are shown in **Figure 2**.

**Figure 2.**
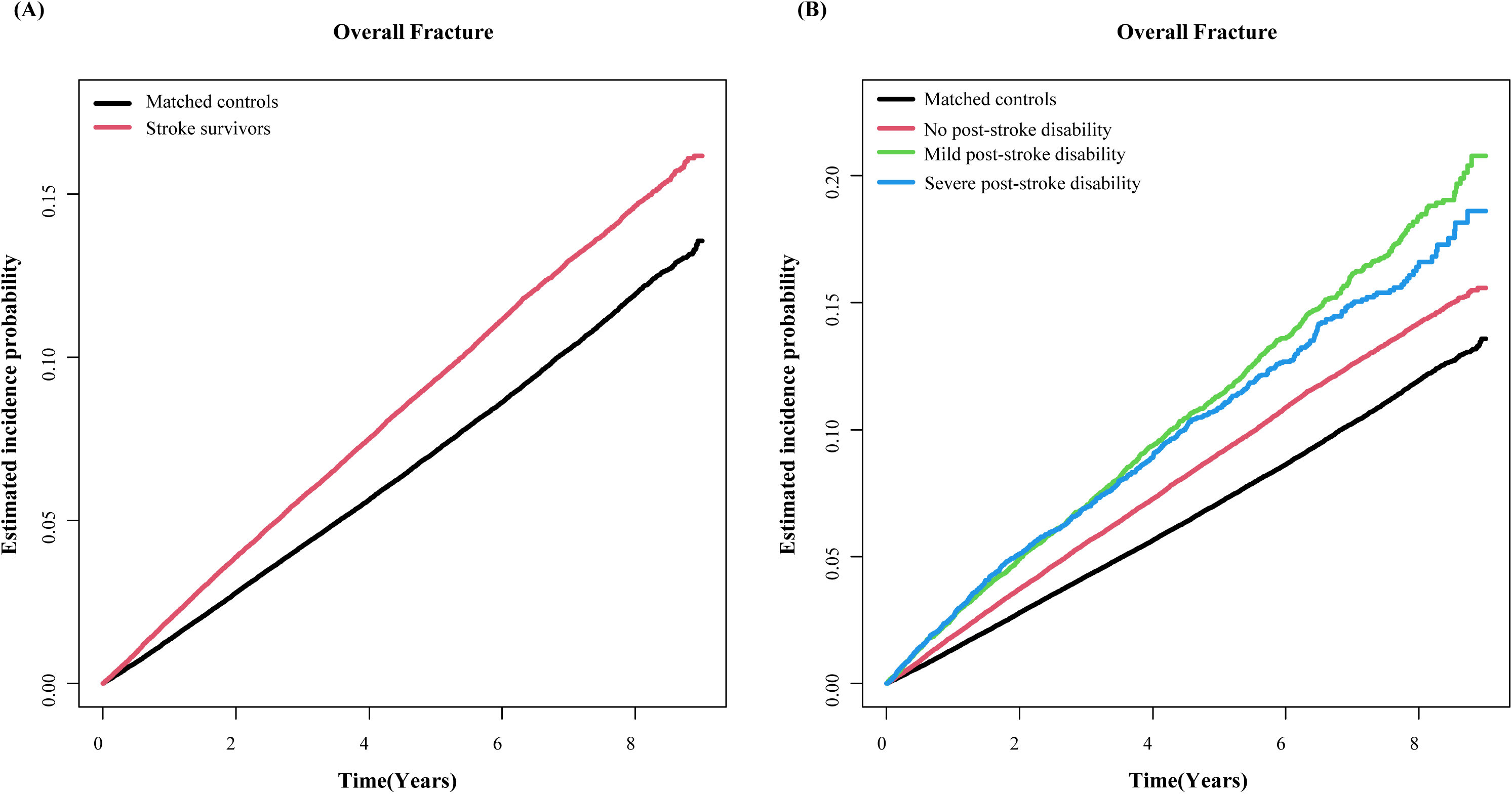
Kaplan-Meier curves displaying the estimated incidence probability of overall fracture A, Risk of overall fracture in stroke survivors and control group; B, Risk of overall fracture in stroke by severity of disability

Stroke survivors had a higher risk of overall fracture (aHR 1.40, 95% CI 1.37-1.43) than the comparison group after adjusting for potential confounders (**Table 2**). Stroke survivors with post-stroke disability had an increased risk of overall fractures (aHR 1.69, 95% CI 1.62-1.76). Stroke survivors without post-stroke disability also had a higher risk of overall fractures compared to the comparison group (aHR 1.36, 95% CI 1.33-1.39).

**Table 2.** Fracture risks among stroke survivors compared to matched controls and according to post-stroke disability.

|  | Number | Event number<br>(n) | IR per 1,000 person-<br>years | Model 1 (Crude)<br>HR (95% CI) | Model 2<br>aHR (95% CI) |
| --- | --- | --- | --- | --- | --- |
| Overall fracture |  |  |  |  |  |
| Matched comparison group | 322,161 | 20,398 | 15.1 | 1 (ref.) | 1 (ref.) |
| Stroke survivors | 223,358 | 16,344 | 19.7 | 1.31 (1.28, 1.34) | 1.40 (1.37, 1.43) |
| No post-stroke disability | 196,426 | 13,885 | 19.0 | 1.27 (1.24, 1.30) | 1.36 (1.33, 1.39) |
| Post-stroke Disability | 26,932 | 2,459 | 24.4 | 1.63 (1.56, 1.70) | 1.69 (1.62, 1.76) |
| Mild post-stroke disability | 17,544 | 1,659 | 24.8 | 1.65 (1.57, 1.74) | 1.67 (1.58, 1.75) |
| Severe post-stroke disability | 9,388 | 800 | 23.6 | 1.57 (1.47, 1.69) | 1.73 (1.61, 1.86) |
| Vertebral fracture |  |  |  |  |  |
| Matched comparison group | 322,161 | 9,797 | 7.1 | 1 (ref.) | 1 (ref.) |
| Stroke survivors | 223,358 | 6,952 | 8.1 | 1.16 (1.12, 1.19) | 1.29 (1.25, 1.34) |
| No post-stroke disability | 196,426 | 6,009 | 8.0 | 1.14 (1.11, 1.18) | 1.28 (1.24, 1.33) |
| Post-stroke Disability | 26,932 | 943 | 9.0 | 1.28 (1.20, 1.37) | 1.39 (1.29, 1.48) |
| Mild post-stroke disability | 17,544 | 691 | 9.9 | 1.42 (1.31, 1.53) | 1.48 (1.36, 1.60) |
| Severe post-stroke disability | 9,388 | 252 | 7.1 | 1.01 (0.89, 1.15) | 1.19 (1.05, 1.35) |
| Hip fracture |  |  |  |  |  |
| Matched comparison group | 322,161 | 3,102 | 2.2 | 1 (ref.) | 1 (ref.) |
| Stroke survivors | 223,358 | 4,027 | 4.7 | 2.14 (2.04, 2.24) | 2.42 (2.30, 2.55) |
| No post-stroke disability | 196,426 | 3,223 | 4.2 | 1.95 (1.86, 2.05) | 2.22 (2.10, 2.35) |
| Post-stroke Disability | 26,932 | 804 | 7.6 | 3.50 (3.23, 3.78) | 3.83 (3.54, 4.15) |
| Mild post-stroke disability | 17,544 | 488 | 7.0 | 3.18 (2.89, 3.50) | 3.38 (3.06, 3.73) |
| Severe post-stroke disability | 9,388 | 316 | 9.0 | 4.13 (3.68, 4.64) | 4.82 (4.28, 5.42) |
| Other fracture |  |  |  |  |  |
| Matched comparison group | 322,161 | 8,933 | 6.5 | 1 (ref.) | 1 (ref.) |
| Stroke survivors | 223,358 | 6,609 | 7.7 | 1.20 (1.16, 1.24) | 1.19 (1.15, 1.23) |
| No post-stroke disability | 196,426 | 5691 | 7.6 | 1.17 (1.14, 1.21) | 1.16 (1.12, 1.21) |
| Post-stroke Disability | 26,932 | 918 | 8.8 | 1.36 (1.27, 1.45) | 1.36 (1.27, 1.45) |
| Mild post-stroke disability | 17,544 | 635 | 9.1 | 1.42 (1.31, 1.53) | 1.39 (1.28, 1.51) |
| Severe post-stroke disability | 9,388 | 283 | 8.0 | 1.24 (1.10, 1.40) | 1.29 (1.14, 1.45) |

IR, incidence rate; HR, hazard ratio; CI, confidence interval
Model 2: adjusted for age, sex, area of residence, insulin, diabetes mellitus, hypertension, dyslipidemia, smoking, alcohol consumption, and body mass index

Stroke survivors exhibited a higher risk of vertebral fracture (aHR 1.29, 95% CI 1.25-1.34) compared to the comparison group (**Table 2**). There was a slight difference in vertebral fracture risk by post-stroke disability status (aHR 1.39, 95% CI 1.29-1.48 vs. aHR 1.28, 95% CI 1.24-1.33). Furthermore, among post-stroke disability, those with mild disability had a higher risk of vertebral fracture (aHR 1.48, 95% CI 1.36-1.60) compared to those with severe disability (aHR 1.19, 95% CI 1.05-1.35).

Stroke survivors had a higher risk of hip fracture (aHR 2.42, 95% CI 2.30-2.55) compared to the comparison group. Furthermore, stroke survivors with post-stroke disability had an even greater risk of hip fracture (aHR 3.83, 95% CI 3.54-4.15) compared to those without post-stroke disability (aHR 2.22, 95% CI 2.10-2.35). Moreover, those with severe disability had a significantly higher risk of hip fracture (aHR 4.82, 95% CI 4.28-5.42) compared to those with mild post-stroke disability (aHR 3.38, 95% CI 3.06-3.73).

### 3. Fracture risk according to type of stroke

We compared the risk of fracture between ischemic and hemorrhagic stroke after stratifying by post-stroke disability status (**Table 3**). No significant difference was found in the risk of overall fracture between the ischemic and hemorrhagic types: 1.36 (1.33-1.40) vs. 1.33 (1.28-1.39) in survivors without post-stroke disability and 1.67 (1.59–1.75) vs. 1.76 (1.62-1.92) with post-stroke disability. However, the risk for vertebral fractures was higher in ischemic stroke survivors compared to hemorrhagic stroke survivors in both strata: without disability (aHR 1.32, 95% CI 1.27-1.36; aHR 1.14, 95% CI 1.07-1.22, respectively) and with disability (aHR 1.46, 95% CI 1.35-1.57; aHR 1.13, 95% CI 0.97-1.33, respectively). The risk of hip fracture was higher for hemorrhagic stroke survivors than ischemic stroke survivors in only those with post-stroke disability (aHR 5.34, 95% CI 4.62-6.16; aHR 3.48, 95% CI 3.18-3.81, respectively).

**Table 3.**
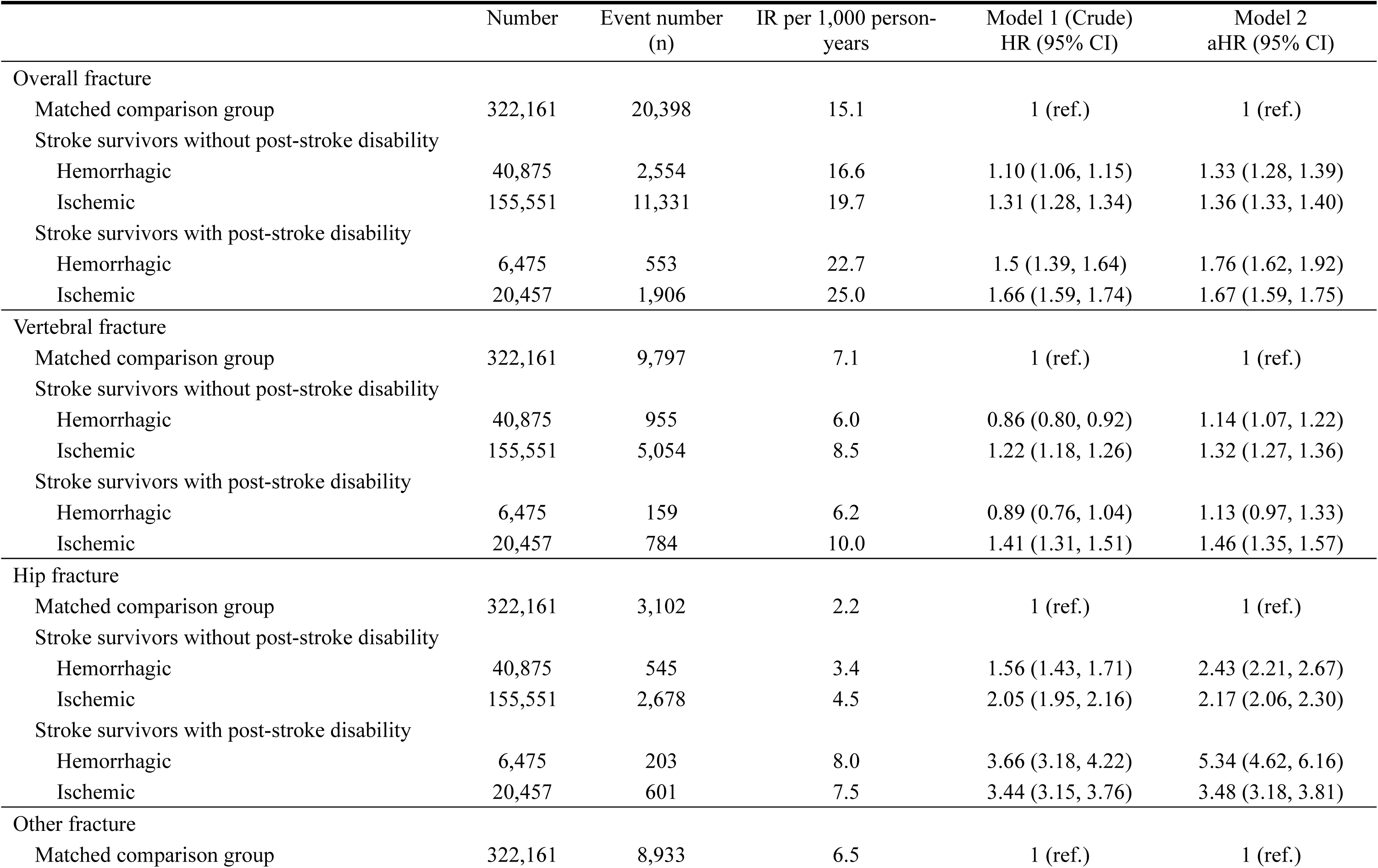
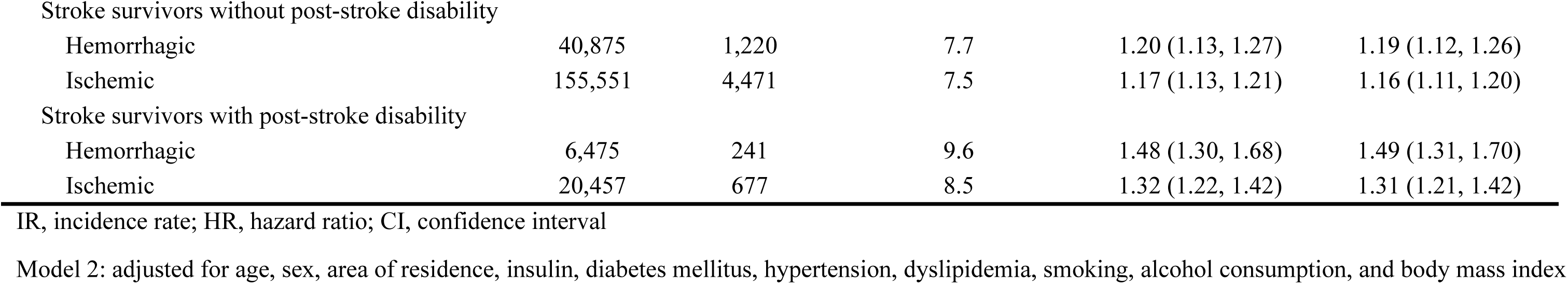
Fracture risks among stroke survivors compared to matched controls and according to type of stroke.

|  | Number | Event number<br>(n) | IR per 1,000 person-<br>years | Model 1 (Crude)<br>HR (95% CI) | Model 2<br>aHR (95% CI) |
| --- | --- | --- | --- | --- | --- |
| Overall fracture |  |  |  |  |  |
| Matched comparison group | 322,161 | 20,398 | 15.1 | 1 (ref.) | 1 (ref.) |
| Stroke survivors without post-stroke disability |  |  |  |  |  |
| Hemorrhagic | 40,875 | 2,554 | 16.6 | 1.10 (1.06, 1.15) | 1.33 (1.28, 1.39) |
| Ischemic | 155,551 | 11,331 | 19.7 | 1.31 (1.28, 1.34) | 1.36 (1.33, 1.40) |
| Stroke survivors with post-stroke disability |  |  |  |  |  |
| Hemorrhagic | 6,475 | 553 | 22.7 | 1.5 (1.39, 1.64) | 1.76 (1.62, 1.92) |
| Ischemic | 20,457 | 1,906 | 25.0 | 1.66 (1.59, 1.74) | 1.67 (1.59, 1.75) |
| Vertebral fracture |  |  |  |  |  |
| Matched comparison group | 322,161 | 9,797 | 7.1 | 1 (ref.) | 1 (ref.) |
| Stroke survivors without post-stroke disability |  |  |  |  |  |
| Hemorrhagic | 40,875 | 955 | 6.0 | 0.86 (0.80, 0.92) | 1.14 (1.07, 1.22) |
| Ischemic | 155,551 | 5,054 | 8.5 | 1.22 (1.18, 1.26) | 1.32 (1.27, 1.36) |
| Stroke survivors with post-stroke disability |  |  |  |  |  |
| Hemorrhagic | 6,475 | 159 | 6.2 | 0.89 (0.76, 1.04) | 1.13 (0.97, 1.33) |
| Ischemic | 20,457 | 784 | 10.0 | 1.41 (1.31, 1.51) | 1.46 (1.35, 1.57) |
| Hip fracture |  |  |  |  |  |
| Matched comparison group | 322,161 | 3,102 | 2.2 | 1 (ref.) | 1 (ref.) |
| Stroke survivors without post-stroke disability |  |  |  |  |  |
| Hemorrhagic | 40,875 | 545 | 3.4 | 1.56 (1.43, 1.71) | 2.43 (2.21, 2.67) |
| Ischemic | 155,551 | 2,678 | 4.5 | 2.05 (1.95, 2.16) | 2.17 (2.06, 2.30) |
| Stroke survivors with post-stroke disability |  |  |  |  |  |
| Hemorrhagic | 6,475 | 203 | 8.0 | 3.66 (3.18, 4.22) | 5.34 (4.62, 6.16) |
| Ischemic | 20,457 | 601 | 7.5 | 3.44 (3.15, 3.76) | 3.48 (3.18, 3.81) |
| Other fracture |  |  |  |  |  |
| Matched comparison group | 322,161 | 8,933 | 6.5 | 1 (ref.) | 1 (ref.) |
| Stroke survivors without post-stroke disability |  |  |  |  |  |
| Hemorrhagic | 40,875 | 1,220 | 7.7 | 1.20 (1.13, 1.27) | 1.19 (1.12, 1.26) |
| Ischemic | 155,551 | 4,471 | 7.5 | 1.17 (1.13, 1.21) | 1.16 (1.11, 1.20) |
| Stroke survivors with post-stroke disability |  |  |  |  |  |
| Hemorrhagic | 6,475 | 241 | 9.6 | 1.48 (1.30, 1.68) | 1.49 (1.31, 1.70) |
| Ischemic | 20,457 | 677 | 8.5 | 1.32 (1.22, 1.42) | 1.31 (1.21, 1.42) |

IR, incidence rate; HR, hazard ratio; CI, confidence interval
Model 2: adjusted for age, sex, area of residence, insulin, diabetes mellitus, hypertension, dyslipidemia, smoking, alcohol consumption, and body mass index

## Discussion

In our large-scale study, stroke survivors had a 1.40 times increased risk of overall fracture compared to the comparison group after adjusting for potential confounders. Among the fracture sites, hip fractures showed the largest risk among stroke survivors (2.42 times) compared to vertebral or other fractures (1.29 and 1.19 times). Stroke survivors with post-stroke disability showed a higher risk than those who did not have post-stroke disability, and severe disability was associated with an even higher hip fracture risk (4.82 times). There was no difference in fracture risk between ischemic and hemorrhagic stroke when post-stroke disability status was accounted for, except for vertebral fractures which showed higher risk in ischemic stroke survivors.

The increased risk of fractures in stroke survivors can be attributed to decreased bone mineral density (BMD) and increased susceptibility to falls following stroke. Bone loss after stroke is presumed to be primarily due to immobilization and increased bone turnover. When the skeleton is not subjected to enough mechanical loading, osteoblast activity decreases, leading to reduced bone synthesis^25^. In a previous study, BMD in the paretic lower limb decreased by 10% among non-ambulatory patients, but only 3% among ambulatory patients^26^. Stroke survivors, even those with mild symptoms are reported to have reduced physical activity^27^. Post-stroke falls typically occur during mobilization^28^, and stroke survivors face an increased risk during basic activities like transferring to and from the bed, chair, and toilet^29^.

Our study found that stroke survivors had a higher risk of both hip and vertebral fractures compared to the control group, but the risk was higher for hip vs. vertebral fracture. As most hip fracture are caused by falls^30^, the increased occurrence of hip fractures in the stroke group could potentially be attributed to falls resulting from diminished balance^6^. Also, the decreased mobility observed in stroke patients may lead to decreased BMD in the hip and subsequent increase in hip fracture risk because the hip bone is particularly sensitive to weight-bearing^26^. In contrast, vertebral fractures are commonly associated with osteoporosis (i.e. compression fracture)^31^ but less frequently associated with falls compared to hip fractures^32^.

Prior studies have shown that there is a significant loss of total body BMD, including hip, but not in vertebrae after a severe stroke^33^. This may be because those with severe disability spend a significant amount of time lying down^5^, leading to reduced weight-bearing activity, which can contribute to greater hip BMD loss^26^. Meanwhile, those with mild disability are able to perform daily activities independently and may require only occasional assistance from others^23^; hence, they have a relatively lower risk of falls and are likely to remain active, and thus are at higher risk of vertebral fractures, which are attributed more to osteoporosis compared to hip fractures^31^. In addition, vertebral fractures were identified through claims data in our study, requiring visit to a hospital. Stroke survivors with mild disability may be more susceptible to experiencing vertebral fracture symptoms compared to those with severe disability, and make hospital visits to investigate the cause and be diagnosed with vertebral fractures. In contrast, stroke survivors with severe disability who have limited access to health care may be less likely to seek medical care for minor injuries such as vertebral fractures. People with functional or mental disabilities have difficulty accessing healthcare^34^.

There was no significant difference in the overall fracture risk between ischemic and hemorrhagic stroke. This is consistent with the results of a recent longitudinal study in Taiwan, which reported that the overall fracture risk did not differ between ischemic and hemorrhagic stroke as the reference (aHR 1.08, 95% CI 0.92-1.16)^7^. However, when assessing site-specific fractures, the risk of vertebral fracture was slightly higher in ischemic stroke survivors with post-stroke disability and without post-stroke disability, while the risk of hip fracture was higher in hemorrhagic stroke survivors with post-stroke disability. This finding aligns with a Taiwanese study, where hemorrhagic stroke was associated with higher risk of hip fracture (aHR 2.34, 95% CI 1.33-4.11) compared with ischemic stroke (aHR 1.60, 95% CI 1.36-1.89)^18^. The underlying reasons for these differential risks of vertebral and hip fractures in ischemic and hemorrhagic stroke survivors remain unclear. However, previous research suggests that, compared with ischemic stroke, hemorrhagic strokes are more severe^35^, and thus greater impaired motor function and bone loss due to immobilization may account for the higher risk of hip fracture in hemorrhagic stroke with post-stroke disability^18^.

Our study has important clinical implications. Delayed rehabilitation of stroke survivors is a risk factor for fracture^36^. Moreover, fractures can result in further functional decline, disrupt stroke rehabilitation, and elevate the risk of subsequent stroke^37,38^. The American Heart Association/American Stroke Association and guidelines for adult stroke rehabilitation and recovery recommend a formal fall prevention program during hospitalization for stroke survivors, and that stroke patients discharged to the community participate in exercise programs with balance training to reduce falls^39^. A previous study demonstrated reduced fracture risk with regular exercise following acute stroke^40^. In addition, it is necessary to conduct examinations and implement preventive treatments for increased bone resorption, which appears to play a significant role in osteoporosis among patients who have experienced prolonged bedridden periods due to immobility^41^. Our study findings highlight the importance of fracture risk prevention among stroke survivors, which may be accomplished by not only fall risk assessments, exercise, and balance training programs^39^, but also by supplementation of calcium and vitamin D^42^ and osteoporosis medications^43^.

This study possesses several strengths, including the utilization of a representative sample obtained from a national database. The substantial number of stroke cases enabled a comprehensive examination of the impact of stroke severity, an aspect often overlooked in prior research. Moreover, stroke severity was defined using a disability grade determined by the Modified Barthel Index, which was validated by specialists. Additionally, the availability of diverse demographic and clinical information facilitated adjustment for confounders in our Cox regression analysis, enhancing the robustness of our findings.

Despite the strength and clinical implication of our study, there are certain limitations to acknowledge. First, the analysis did not consider the specific lesion site of stroke as the ICD-10 codes or KNDRS used in this study do not distinguish different lesion sites. Stroke location affects clinical outcomes^44^, and future studies should identify the specific lesion site to help identify stroke survivors more vulnerable to fracture risk. Second, given that the definition of fractures relied on claims data using ICD-10 codes, there is a possibility of under detection of fractures, especially for vertebral fractures. Third, because we used an established public database, we were unable to analyze several factors, including the cause of fractures, family history of fracture, BMD, balance or cognitive impairment, or antithrombotic medication status, which may affect fracture risk. Future research should focus on identifying the impact of these factors on fracture risk among stroke survivors. Furthermore, our study participants were restricted to those who received health screening examinations, potentially representing a cohort that is healthier and more engaged in healthy lifestyles compared to the general population. Finally, the use of a single-country database limits the generalizability of our study to other populations due to differences in medical treatment approaches that may differ by local practice guidelines and healthcare systems, cultural environments, and ethnicity^45^.

In summary, our study showed that stroke survivors had a 1.4-fold increased risk of overall fracture and a 2.4-fold increased risk of hip fracture. These risks were further elevated among individuals with post-stroke disability. When stratifying by post-stroke disability status, no significant difference was observed between ischemic and hemorrhagic stroke for overall fracture risk. Fall prevention and comprehensive bone health management should be prioritized in the care of stroke survivors.

## Data Availability

The database is open to all researchers whose study protocols are approved by the official review committee.

## Acknowledgements

The authors thank Da-Hyeun Lee and Sang-Eun Lee for the graphical abstract.

## Author contributions

D.L. drafted and revised the article. I.Y.C. and D.W.S. interpreted the data and contributed to the study conception and design. K.H. was involved in data acquisition and statistical analysis. W.H.C., J.E.Y., H.L.C., and J.H.P. revised manuscript. All authors read and agreed to the final manuscript.

## Data Availability Statement

The KNHIS database is open to researchers whose study protocols are approved by the official review committee.

## Supplemental Material

Supplementary Methods

Tables S1-S4

